# HIV viral non-suppression and drug resistance among persons who inject drugs on dolutegravir antiretroviral therapy in Kenya

**DOI:** 10.64898/2026.02.26.26347230

**Authors:** Loice W. Mbogo, Ceejay L. Boyce, Betsy Sambai, Stephen E. Hawes, Brandon L. Guthrie, William Sinkele D. Min, David Kimani, Vincent Adhanja, Bhavna H. Chohan, Robert A. Smith, Aliza Monroe-Wise, Esther Gitau, Sarah Masyuko, Vincent C. Marconi, Geoffrey S. Gottlieb, Paul K. Drain, Lisa M. Frenkel, Carey Farquhar DREAM-Kenya

**Affiliations:** University of Washington Global Assistance Program-Kenya, Nairobi, Kenya; Division of Infectious Diseases Emory University School of Medicine and Department of Global Health, Emory University, Atlanta, USA; Center for Global Infectious Disease Research, Seattle Children’s Research Institute, Seattle, USA; Department of Epidemiology, School of Public Health, University of Washington, Seattle, USA; Department of Global Health, University of Washington, Seattle, USA School of Public Health and School of Medicine, University of Washington, Seattle, USA; National HIV and STI Control Programme (NASCOP), Kenya Ministry of Health, Nairobi, Kenya; Support for Addictions Prevention and Treatment in Africa (SAPTA), Nairobi, Kenya; Department of Medicine, Division of Allergy and Infectious Diseases, University of Washington, Seattle, USA; Department of Pediatrics, University of Washington, Seattle, USA; Department of Laboratory Medicine & Pathology, University of Washington, Seattle, USA

**Keywords:** Persons who inject drugs (PWID), dolutegravir, antiretroviral therapy (ART), viral non-suppression, drug resistance, longitudinal cohort study

## Abstract

**Background:** Maintaining viral suppression among people who inject drugs (**PWID**) living with HIV in sub-Saharan Africa remains critical to minimize drug resistance for dolutegravir (**DTG**)-based regimens. We evaluated PWID taking DTG to assess longitudinal rates of viral non-suppression and emergence of drug resistance mutations in Kenya.

**Methods:** We enrolled Kenyan PWID who had transitioned from an efavirenz (**EFV**) based regimen to tenofovir+lamivudine+DTG (**TLD**) ≥6 months prior, and measured plasma HIV RNA viral load (**VL**) every 6 months for 2 years. We used univariable Cox proportional hazards to assess longitudinal risk for viremia (VL >200 copies/ml). Plasma specimens with viremia were genotyped for HIV drug resistance, including minority variants, using a lab-developed PacBio sequencing assay, and referenced by the Stanford HIVdb program.

**Results:** Among 250 participants, 125 were receiving methadone, 199 (79.6) reported heroin use, 70% were male, and median age was 39 years. 194 (77.6%) participants completed all five study visits, 41 (16.4%) were lost to follow-up and 15 (6.0%) died. Across all study visits, 166 (66.0%) of the 250 participants were always suppressed, and 84 (33.6%) were viremic at least once during follow-up, including 8 (3.2%) who were always viremic and 76 (30.4%) who were intermittently suppressed. Living in an improvised shelter or outdoors was significantly associated with a higher risk of viremia (HR=4.35, 95% CI: 1.52–12.53). 93 specimens had drug resistance genotyping, 27 (29%) of which were from participants with incomplete follow-up. NNRTI resistance was frequent (37-41% across visits), whereas major resistance mutations were infrequent to tenofovir (4.3%), lamivudine (7.5%), and DTG (1%, minority variant S153F detected at 1% frequency). Accessory DTG mutations, which do not independently reduce susceptibility, were more common, observed in 41% (38/93) of genotyped specimens, most often T97A, E138K, and L74M.

**Conclusion:** Among PWID living with HIV on TLD in Kenya, one-third had intermittent or sustained viral non-suppression across two years of follow-up. While NNRTI resistance was common, DTG resistance mutations were rare. Improving viral suppression among PWID living with HIV will reduce transmission risks and improve clinical outcomes.

## Introduction

Among people who inject drugs (**PWID**) living with HIV in sub-Saharan Africa, there is a paucity of data on the real-world effectiveness of dolutegravir (**DTG**)-based regimens. PWID encounter numerous challenges in accessing services, such as incarceration, displacement, homelessness, transportation expenses, stigma, and food insecurity, which can complicate HIV care and adherence and may impact outcomes [1]. In a multi-country study, more than half of PWID were not virally suppressed, and behavioral and demographic factors played a major role, suggesting a need to enhance HIV monitoring and care for members of this key population[2].

The World Health Organization (**WHO**) recommends newer integrase strand transfer inhibitors (**INSTI**) in first-line ART regimens like the 3-drug regimen TLD: tenofovir disoproxil fumarate (**TDF**) + lamivudine (**3TC**) + dolutegravir (**DTG**) [3]. A systematic review showed that DTG has high durability, safety, and efficacy [4]. The development and advancement of HIV antiretroviral treatment (**ART**) have brought a shift to more effective, better tolerated, and safer drug options among people with HIV (**PWH**) worldwide [5], [6]. While prevalence is low, recent studies raise concern for increasing INSTI resistance with widespread use [7], [8], [9]. A recent study in sub-Saharan African (**SSA**) found the emergence of major DTG resistance mutations in 2.4% of INSTI-naive patients [10].

In 2017, the Kenyan Ministry of Health National AIDS & STI Control Program rolled out the use of TLD as first-line ART among the general population of PWH[11]. In 2019, TLD was rolled out in drop-in centers serving key populations, including for men who have sex with men, female sex workers, and PWID. In Kenya, HIV prevalence in the general population had reduced to 3% among adults in 2024 [12], due to interventions such as test and treat, assisted partner notification services, reduced pill burden, and better-tolerated treatment [3], [13]. Yet HIV prevalence among PWID remains relatively high at 18.7% despite interventions aimed at reducing HIV transmission among Kenyan PWID [14]. Viral non-suppression is thought to play a role in the high HIV prevalence and potential for ongoing transmission among PWID. A cross-sectional study done in Nairobi and Coastal Kenya among PWID on ART before switching to a DTG-containing regimen showed that 148 (23%) had unsuppressed HIV-1 plasma RNA viral loads (**VL**) defined as >1,000 copies/Ml [15]. Of those, 122 (82.4%) of 148 had HIV-1 polymerase gene sequences available, and 55 (45.1%) had major HIV-drug resistance mutations of non-nucleoside and nucleoside reverse transcriptase inhibitors (**NNRTI** and **NRTI**) [15].

This study was undertaken to evaluate VL and TLD drug resistance trends among PWID living with HIV in Kenya, a key population disproportionately affected by the epidemic. These insights are essential for strengthening HIV programs for PWID on DTG-containing regimens, optimizing treatment outcomes, and advancing progress toward epidemic control in Kenya and globally.

## Methods

### Study Design, Participants, and Procedures

We conducted a prospective cohort study that enrolled participants who were actively receiving services in drop-in centers and methadone clinics in Nairobi and the Kenyan coastal region. Data were collected between May 2021 and September 2023 with baseline, 6, 12, 18, and 24-month follow-up visits. Pre-DTG initiation under Kenya ministry of health guidelines, all individuals aged 15 years were initiated on TDF + 3TC + efavirenz (**EFV**) as first-line treatment [16]. In 2018 WHO recommended switching to TLD as a preferred first-line regimen [17], which Kenya was first in East Africa to implement in the general population [11]. Later in July 2019, Kenya scaled up the use of DTG-based regimens among key populations. All individuals receiving care at methadone clinics at the Ministry of Health center and NSPs site were initiated on DTG regardless of their viral load result, which were assessed every 6 months or annually. Site clinicians referred PWID who had been on DTG for at least 6 months to a trained study health advisor who recruited those who were ≥18 years of age and had injected drugs in the past year. In Nairobi, participants were recruited from the methadone clinic at Ngara Health Centre and from three needle and syringe program (**NSP**) sites managed by a harm reduction organization, Support for Addiction Prevention and Treatment in Africa (**SAPTA**). In the coastal region, participants were recruited from 4 NSP sites, including the Reachout program in Mombasa, the Muslim Education Welfare Association (**MEWA**) sites in Mtwapa and Kilifi, and The Omari Project in Malindi.

At enrollment, participants provided written informed consent, completed a questionnaire, and had blood drawn for HIV VL and genotypic resistance testing. The 6, 12, 18, and 24 month follow-up visits included questionnaires and an additional blood draw for the same assays. All data were entered into structured questionnaires programmed in REDCap®. A laboratory request form was used to document blood collection at each visit.

### Laboratory Methods

HIV VL was measured at each visit (baseline, 6, 12, 18 and 24 months with a window of 90 days) using the Abbott RealTime HIV-1 Viral Load assay performed in the University of Nairobi’s Molecular Medicine and Infectious Diseases laboratory. HIV VL results were provided to the clinic for participants’ clinical care and ART management.

Viremic specimens, defined as HIV VL >200 copies/ml, were genotyped for HIV drug resistance mutations prospectively at Seattle Children’s Research Institute using a laboratory-developed assay for the PacBio sequencing platform. The in-house assay amplifies a ∼3kb region of HIV *pol* encoding protease, reverse transcriptase, and integrase (codons PR19-IN270) as previously described [18]. If PacBio sequencing was not successful, typically due to lower VL, Sanger sequencing was performed on amplicons (codons PR4-RT251 and IN44-270) generated by nested RT-PCR as described [18]. A subset of participants who had been enrolled in an earlier study, called the SHARP study, [15, 19] had viremic samples available from visits prior to DTG initiation; these were sequenced to determine if drug resistance before starting DTG was associated with viremia on TLD. Sequencing data can be accessed via BioProject PRJNA1256474. The prevalence of drug resistance to NRTI, NNRTI, and INSTI were assessed using the Stanford HIVdb program (https://hivdb.stanford.edu) with resistance to NRTI or NNRTI defined as a cumulative mutation score ≥30 and “major” DTG resistance defined as a mutation score >10.

### Ethical approvals

Ethical approvals were obtained from Kenyatta National Hospital/University of Nairobi Ethical and Scientific Review Committee in Kenya, and the Seattle Children’s Institutional Review Board in the USA. Participants were requested to provide written informed consent in Swahili or English. PWID may sometimes be under the influence and cognitively impaired, thus unable to make sound judgments. Study staff were trained to reschedule participants who could not comprehend information and to enroll only when they could make informed decisions about participation. All study sites had dedicated research rooms to offer privacy during study procedures.

### Data analysis

The IRIS Biometric system that generated random numbers was used to select a unique identifier, enabling the study team to confirm participant identity every time they presented for study visits [20]. Biometric unique identifiers prevented duplication in participant enrolment and overlap within sites during follow-ups [21].

Descriptive statistics were reported using medians and interquartile ranges (**IQR**) (***Table 1***) for continuous variables and percentages for categorical variables. Fisher’s exact test was used to assess the association between pre-treatment drug resistance and HIV VL non-suppression (**VLNS**) for the subset of participants with pre-DTG genotypes and Cox regression survival analysis was used to determine the factors associated with viremia.

**Table 1.** Sociodemographic characteristics of 250 participating PWID living with HIV.

|  |  | **Mean (SD) or number**<br>**(percent)**<br>**Total N=250** |
| --- | --- | --- |
| Age |  | 38.6 (7.9%) |
| Sex | Female | 75 (30.0%) |
| Marital status | Divorced/separated/widowed | 101 (40.4%) |
|  | Married/partnered | 87 (34.8%) |
|  | Single | 62 (24.8%) |
| Education | None/Primary | 170 (68.0%) |
|  | Secondary/College | 80 (32.0%) |
| Living condition | Home/Relative | 231 (92.4%) |
|  | Improvised shelter/Open | 19 (7.6%) |
| Years injecting |  | 7.6 (5.8%) |
| ***Substance use in the last 12months*** |  |  |
| Heroin | Yes | 199 (79.6%) |
| Alcohol | Yes | 57 (22.8%) |
| Valium | Yes | 13 (5.2%) |
| Artane | Yes | 4 (1.6%) |
| Rohypnol | Yes | 16 (6.4%) |
| Khat | Yes | 43 (17.2%) |
| Cocaine | Yes | 21 (8.4%) |
| Jail ever | Yes | 185 (74.0%) |
| WHO stage | 1 | 138 (55.2%) |
|  | 2 | 23 (9.2%) |
|  | 3 | 89 (35.6%) |
| Methadone | Yes | 125 (50%) |

Abbreviations: Interquartile range (IQR) and Standard deviation (SD) used on the table

The outcome variable was defined as the time to first episode of viremia >200 copies/ml (e.g., loss of VL suppression). Event predictors were adjusted by characteristics such age, sex, living conditions, duration of substance use, marital status, education, heroin use, alcohol use, methadone participation, and if ever jailed. We did not determine factors associated with drug resistance to dolutegravir because this resistance was low at <2%. Participants who missed some visits were censored if the event did not occur within the 24-month follow-up period. Data analyses were performed using the R statistical program (version 4.1.2) and the significance level was set at alpha= 0.05.

## Results

### Participant characteristics

Median age for the 250 participants was 39 years (IQR: 33-44); they were predominantly male 175 (70%), and most 170 (68%) had attained primary education or less (***Table 1***). Participants had injected drugs for a median of 5 years (IQR: 3-10), 125 (50%) were on methadone and 199 (79.6%) reported use of heroin, indicating that some of the participants were using two. Nineteen (7.6%) were living in an improvised shelter or in the open, and 185 (74%) reported ever having been incarcerated. The drugs that were used most frequently in the past month were heroin, used by 199 (79.6%), and alcohol, used by 57 (22.8%). More than half of the participants were in WHO stage 1 of HIV infection and 203 (81.2%) were virally suppressed at enrollment. The proportion of participants who were virally suppressed remained relatively constant over time, ranging from 81% to 85% (***Table 2***). Among the 250 participants across all of their study visits, 166 (66.0%) were always suppressed, and 84 (33.6%) were viremic at least once during follow-up, including 8 (3.2%) who were always viremic and 76 (30.4%) who were intermittently suppressed. Of the 250 enrolled participants, 194 (77.6%) had complete 2-year follow-up (n=5 visits). Of these, 127 (65.5%) were always suppressed, and 67 (34.5%) were viremic at least once during follow-up, including 3 (4.5%) of whom were always viremic and 64 (95.5%) of whom were intermittently viremic. A total of 41 (16.4%) participants were lost to follow-up, of which 15 (6.0%) died. An additional 27 participants with incomplete follow-up experienced viremia at least once during follow-up, for a total of n=94 (99%) participants, and the median HIV VL when viremic was 16,654 copies/ml (IQR: 1,759-101,881 c/mL) across all study visits.

**Table 2.** Proportion of participants with HIV viral suppression at each visit.

|  | Enrollment<br>n/250 (%) | Month 6<br>n/231 (%) | Month 12<br>n/223 (%) | Month 18<br>n/217 (%) | Month 24<br>n/205 (%) |
| --- | --- | --- | --- | --- | --- |
| **Suppressed**<br>**(<200 c/mL)** | 203 (81%) | 185 (80%) | 183 (82%) | 184 (85%) | 166 (81%) |
| **Viral non-**<br>**suppression**<br>**(≥200 c/mL)** | 47 (19%) | 46 (20%) | 40 (18%) | 33 (15%) | 39 (19%) |

### TLD drug resistance and virologic outcome

Of the 250 participants enrolled in this study, 61 (24.4%) had pre-DTG genotypic data available from their enrollment visit in the SHARP study [15], [22]. We found that having a pre-treatment drug resistance mutation was not associated with viral non-suppression at one or more visits on DTG-ART (59% vs 54%, p=0.8) (***Table 3***). Moreover, pre-treatment drug resistance specifically to the NRTI backbone in DTG-ART (TDF/3TC) was not associated with viral non-suppression on DTG-ART (75% vs 54%, p=0.6).

**Table 3.** Viral suppression status and presence of pre-treatment drug resistance to DTG-ART among 122 participants who had HIV-I polymerase gene sequences available.

|  | Any Drug<br>Resistance at<br>SHARP<br>Enrollment<br>n (%)<br>(N=22) | Wild-Type at<br>SHARP Enrollment<br>n (%)<br>(N=39) | PDR to<br>TDF/3TC<br>at SHARP<br>Enrollment<br>n (%)<br>(N=4) | No PDR to<br>TDF/3TC at<br>SHARP<br>Enrollment<br>n (%)<br>(N=57) |
| --- | --- | --- | --- | --- |
| **Always**<br>**Suppressed** | 9 (41%) | 18 (46%) | 1 (25%) | 26 (46%) |
| **≥1 Viremic Visit** | 13 (59%) | 21 (54%) | 3 (75%) | 31 (54%) |
| **Intermittently**<br>**changing** | 7 | 13 | 2 | 18 |
| **Always Non-**<br>**suppressed** | 6 | 8 | 1 | 13 |

### Prevalence of HIV drug resistance

During follow-up, a total of 94 specimens from participants who experienced viremia at different time points, including 27 with incomplete follow up and 67 who completed all five visits, were selected for genotyping. Genotyping was successful for 93 (99%) of these specimens.

Resistance to NNRTIs was common (∼37-41% of viremic specimens), but major drug resistance mutations associated with the drugs in their DTG-based regimen were relatively rare (***Table 4***). Mutations associated with resistance to TDF and 3TC were found in 7/93 (8%) specimens and major DTG resistance was only observed as minority variants in 1/93 (1%) specimens: S153F mutation was detected in 2/157 (1%) viral templates sequenced. Mutations classified as “accessory resistance mutations” for DTG, which alone do not confer reduced susceptibility to DTG, were observed at higher frequency in 38/93 (41%) participants with the most common mutations being T97A (n=7), E138K (n=7), and L74M (n=6).

**Table 4.** Genotype prevalence of HIV drug resistance at visits with viremia.

|  | Month 0<br>(N=47) | Month 6<br>(N=46) | Month 12<br>(N=40) | Month 18<br>(N=33) | Month 24<br>(N=39) |
| --- | --- | --- | --- | --- | --- |
| **Number (%) with Genotyping Data** | 46 (98%) | 46 (100%) | 40 (100%) | 32 (97%) | 38 (97%) |
| **Wild-type** | 28 (61%) | 28 (61%) | 25 (63%) | 19 (59%) | 23 (61%) |
| **Any Drug Resistance Mutation(s)** | 18 (39%) | 18 (39%) | 15 (37%) | 13 (41%) | 15 (39%) |
| *TDF/3TC Drug Resistance* | 5 (11%) | 5 (11%) | 4 (10%) | 1 (3%) | 4 (10%) |
| *DTG Drug Resistance* | 0 (0%) | 0 (0%) | 0 (0%) | 1 (3%) | 0 (0%) |
| *DTG Accessory Drug Resistance Mutation(s)* | 13 (28%) | 14 (30%) | 11 (28%) | 11 (33%) | 14 (36%) |

Abbreviations: Tenofovir disoproxil fumarate (TDF), Lamivudine (3TC,) Dolutegravir (DTG)

### Factors associated with time to viremia

To determine factors associated with viremia, we evaluated 203 participants who were suppressed at the baseline and had 2 or more follow-up visits. In the univariate Cox regression model, living in an improvised shelter was associated with becoming viremic (HR=4.36, 95% CI: 1.52,12.53, p=0.006) (***Table 5***). In the adjusted model, there was a trend for living in an improvised shelter to be associated with shorter time to viremia, but this did not attain statistical significance (HR=3.04, 95% CI: 0.96, 9.60). No other variables were associated with time to viremia. We carried out a secondary analysis, censoring any missingness of HIV VL and results did not change substantially.

**Table 5.** Cox regression analysis to determine factors associated with viral non-suppression.

|  |  | **Unadjusted Hazard Ratio (HR)**<br>**(95% CI)** | **Adjusted (aHR) (95%**<br>**CI)** |
| --- | --- | --- | --- |
| Age |  | 0.98 (0.93-1.02), p=0.27 | 0.96 (0.91-1.01), p=0.12) |
| Sex | female | Reference | - |
|  | male | 1.51 (0.69-3.31), p=0.30) | 1.89 (0.81-4.43), p=0.14) |
| Marital status | Divorced/separated/widowed | Reference | - |
|  | Married/partnered | 0.80 (0.40-1.60), p=0.52) | 0.79 (0.37-1.67), p=0.53) |
|  | Single | 1.09 (0.45-2.65), p=0.86) | 0.73 (0.26-2.02), p=0.55) |
| Education | None/Primary | - | - |
|  | Secondary/College | 0.53 (0.23-1.21), p=0.13) | 0.57 (0.25-1.30), p=0.18) |
| Living condition | Home/Relative | - | - |
|  | Improvised shelter/Open | **4.36 (1.52-12.53), p=0.006)\*** | 3.04 (0.96-9.60), p=0.06) |
| Years injecting | <5 | - | - |
|  | >five | 0.71 (0.37-1.36), p=0.30) | 0.74 (0.36-1.51), p=0.40) |
| Heroin use in the past month | No | - | - |
|  | Yes | 4.76 (0.65-34.74), p=0.12) | 4.46 (0.60-32.91), p=0.14) |
| Alcohol | No | - | - |
|  | Yes | 0.64 (0.34-1.22), p=0.17) | 0.72 (0.37-1.40), p=0.34) |
| Valium | No | - | - |
|  | Yes | 0.28 (0.04-2.06), p=0.21) | - |
| Rohypnol | No | - | - |
|  | Yes | 0.47 (0.17-1.34), p=0.16) | - |
| Khat | No | - | - |
|  | Yes | 0.98 (0.52-1.83), p=0.94) | - |
| Cocaine | No | - | - |
|  | Yes | 1.90 (0.83-4.33), p=0.13) | - |
| Jail ever | No | - | - |
|  | Yes | 1.53 (0.64-3.66), p=0.34) | - |
| WHO stage | One | - | - |
|  | Two | 1.13 (0.58-2.22), p=0.71) | - |
|  | Three | 1.01 (0.24-4.31), p=0.99) | - |
| Methadone participation | Not on Methadone | - | - |
|  | On Methadone | 0.71 (0.37-1.35), p=0.30) | 0.67 (0.34-1.33), p=0.25) |

**Abbreviations:** Hazard Ratio (HR), Adjusted Hazard Ratio (aHR), or Confidence intervals (CI) .
**Adjusted for** age, sex, marital status, education, living conditions, years of injecting, heroin use in the past one year, alcohol use and methadone participation.

## Discussion

In this study, we closely monitored virologic suppression and drug resistance patterns over a 24-month period in a large cohort of 250 PWID living with HIV in Kenya and taking TLD. Our primary findings were that while a large proportion had either persistent or intermittent viremia, drug resistance was low in this cohort. Of those who developed viremia during the study, being unhoused was the only risk factor we identified. Interestingly, having NRTI or NNRTI drug resistance prior to starting TLD was not a risk factor for viremia or development of drug resistance among a subset of participants with pre-DTG genotype data, confirming findings of the large, randomized NADIA trial[23].

Over the two-year follow-up period, participants in this study fell into three categories of virologic outcomes: consistently suppressed, consistently viremic, and intermittently viremic. The majority of participants remained consistently suppressed, while a smaller proportion were persistently viremic or experienced intermittent viremia. That most participants in our cohort receiving a DTG-containing ART regimen (i.e., TLD) remained consistently virally suppressed over time aligns with evidence from a multinational African study that demonstrated improved viral suppression rates over time among DTG naïve individuals following a switch to DTG-based ART [24].

Yet, while the majority were suppressed and there was little drug resistance, prevalence of viremia among PWID in this longitudinal study was approximately twice as high as that reported in the general population of PWH in Kenya, with viremia detected in 15-20% of participants. One reason we may have seen such a large number with intermittent rather than persistent viremia is that peer educators were mobilized to trace participants with viremia and provide adherence counseling, working closely with clinicians to ensure re-engagement in care and subsequent suppression. This is particularly important in the current context, as the planned withdrawal of support from the U.S President’s Emergency Plan for AIDS Relief (PEPFAR) in 2025 threatens to disrupt the systems that enable continuity of HIV care among key populations. This suggests that close monitoring of this population could be beneficial, with more frequent, biannual HIV VL assessments rather than the standard once a year or every two years.

In our study, HIV viremia among PWID was associated with living in an improvised shelter or out in the open, suggesting that PWID with housing instability are at high risk for poor outcomes. We initially hypothesized that being on methadone would provide some level of protection since studies have shown that methadone use is associated with viral suppression [25](Mbogo et al., 2021). Notably, many of those on methadone continued using heroin. Other studies have highlighted social determinants including stigma, homelessness, and incarceration as key contributors to virologic failure [26], [27], [28]. We found no difference in the time to viremia among those on methadone when compared to those not on methadone, which may be explained by the fact that all the participants, regardless of their methadone status, had regular visits and were closely monitored throughout the study.

DTG drug resistance was infrequent among the PWID over a two-year follow-up period in this cohort. However, exposure to DTG was limited since it had only recently been introduced into PWID cohorts and follow-up was short at only two years. Another factor to consider is that the viremia observed in this study population had relatively high viral loads which suggests participants had long periods of complete ART non-adherence which may be less likely to select resistance given the lack of drug pressure on the virus to select mutations, especially for a single tablet regimen composed of antiretrovirals with similar half-lives. In addition, drug resistant variants generally have relatively poor replication capacities and during period of non-adherence are out-grown by wild-type variants.

While resistance was uncommon in our cohort, the prevalence of DTG resistance is increasing. A predictive DTG resistance analysis model in South Africa estimates an increase in acquired dolutegravir resistance from 18.5% to 41.7% between 2020-2035 [8], [29] although the differences in these estimates are in part due to the mutations used to define DTG resistance. Because E138K does not independently confer DTG resistance by the Stanford scoring, this mutation alone was not considered to confer resistance in some studies, while it was in others [8]. Currently, evidence on drug resistance to DTG-containing regimens among the PWID in Sub-Saharan African (**SSA**) remains limited compared to the general population of PWH, raising questions about the prevalence resistance in viremic individuals [10], [30]. Extended longitudinal studies of HIV drug resistance with more frequent VL monitoring are needed to better characterize the emergence and dynamics of resistance-associated mutations among PWID and their sexual partners.

## Data Availability

Data availability All data is stored under the protection and guidelines of the University of Washington in Seattle, specifically in a dedicated OneDrive.

## Authors’ Contribution

CF, LF LWM, CLB, PKD, GSG, AM, SEH, VM, BLG, SM, contributed to the research concept, study implementation, and data analysis. BS, DK, and VA were involved in data collection and further data analysis. WS, EG, SM, and BC supported the data analysis process. LWM wrote the initial draft of the manuscript, CF, CB, SH revised subsequent drafts and all authors provided input on the final version. All authors accessed the data and approved the manuscript before submission.

## Acknowledgment

We extend gratitude to all the participants and staff in Kenya who contributed to the study. We extend our appreciation to all NSP sites (MEWA, SAPTA, TOP, and Reachout) for their valuable support.

